# DDR-Augmented-Artifacts: Synthetic Artifact Overlays for Robust Diabetic Retinopathy Models

**DOI:** 10.1101/2025.09.25.25336606

**Authors:** Shubham Aggarwal

## Abstract

Deep learning models for the screening of diabetic retinopathy (DR) have achieved near-human performance on benchmark datasets, but their performance deteriorates in real-world settings due to imaging artifacts such as glare, blur, and reflections. Current public datasets such as DDR contain high-quality fundus images, but they lack the variability and imperfections seen in handheld fundus photography. This mismatch results in models that fail in practice, particularly in low-resource environments where handheld cameras are widely deployed.

We introduce **DDR-Augmented-Artifacts**, an artifact-augmented extension of the DDR dataset that simulates realistic reflection artifacts via patch-based Poisson blending. Unlike prior datasets that exclude noisy images, our dataset explicitly models these challenges, allowing researchers to benchmark and train models that are robust to real-world noise. The dataset, augmentation scripts, and a sample demonstration model are publicly available at:

- GitHub: https://github.com/Shubham2376G/DR_Artifacts
- Hugging Face: https://huggingface.co/datasets/shubham212/DR_Artifacts

## 1 Introduction

Diabetic Retinopathy (DR) is one of the leading causes of preventable blindness. Deep learning has enabled automated DR severity classification into *referable* vs. *non-referable* DR, with performance rivaling that of expert ophthalmologists. However, a major limitation of existing AI solutions is their reliance on clean, high-quality images obtained from tabletop fundus cameras under controlled conditions.

**Table 1:** Reported performance of landmark AI systems for referable DR detection on high-quality tabletop fundus images. These results demonstrate near-expert performance under ideal conditions.

| Study / System | Dataset / Setting | AUC | Sensitivity | Specificity |
| --- | --- | --- | --- | --- |
| Gulshan et al. (2016) [3] | EyePACS (USA) | 0.991 | 97.5% | 93.4% |
|  | Messidor-2 | 0.990 | 96.1% | 93.9% |
| Ting et al. (2017) [4] | Multi-ethnic (Singapore) | 0.936 | 90.5% | 91.6% |
| IDx-DR (FDA cleared) [5] | Clinical tabletop cameras | – | 87.2% | 90.7% |
| EyeArt (FDA cleared) [6] | Clinical tabletop cameras | – | 96.0% | 88.0% |

In real-world screening:

- Patients may not be able to comfortably undergo repeated image captures, making it impractical to discard poor-quality images.
- Retakes increase screening time, cost, and operator workload.
- In rural and low-resource settings, handheld devices are increasingly used, where imaging artifacts are more common.

Thus, models trained only on pristine datasets perform poorly in deployment. We address this gap by releasing DDR-Augmented-Artifacts, a dataset that augments DDR images with realistic reflection artifacts. DDR-Augmented-Artifacts provides a reproducible, artifact-augmented dataset for diabetic retinopathy research. Scripts and an optional demonstration model allow researchers to benchmark robustness under acquisition artifacts.

## 2 The Problem of Reflection Artifacts

Reflection artifacts are bright, shiny regions that appear in handheld fundus photographs. They are caused by the following:

- Misalignment of illumination and optical axis.
- Specular reflection from the inner limiting membrane or blood vessels.
- Instability of handheld devices compared to fixed tabletop cameras.

**Figure 1.**
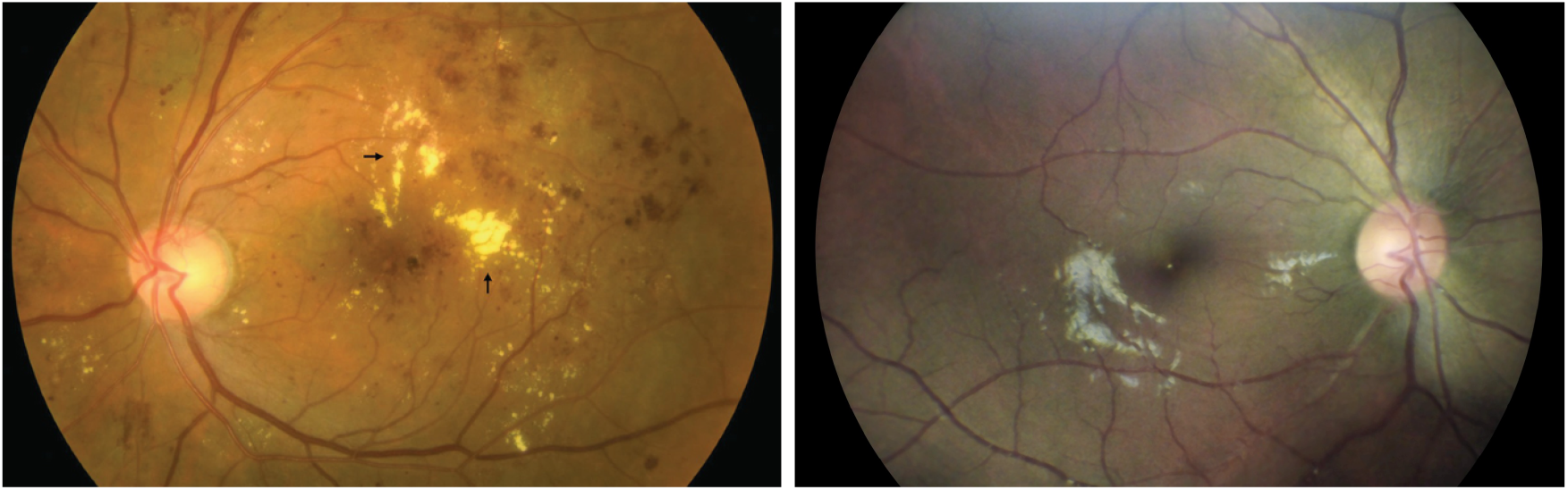
Left: hard exudates in a retinal fundus image. Right: retinal fundus image with reflection artifact (white shiny areas).

### 2.1 Clinical Risk

These artifacts are visually similar to **hard and soft exudates**, key indicators of DR. Misclassification risks include:

- **False Positives:** Reflection misclassified as exudates → overestimation of severity.
- **False Negatives:** True lesions hidden under reflection → underestimation of severity.

This can lead to unnecessary referrals, missed diagnoses, and erosion of clinician trust in AI systems.

## 3 Comparison: Tabletop vs. Handheld Imaging

**Table 2:**
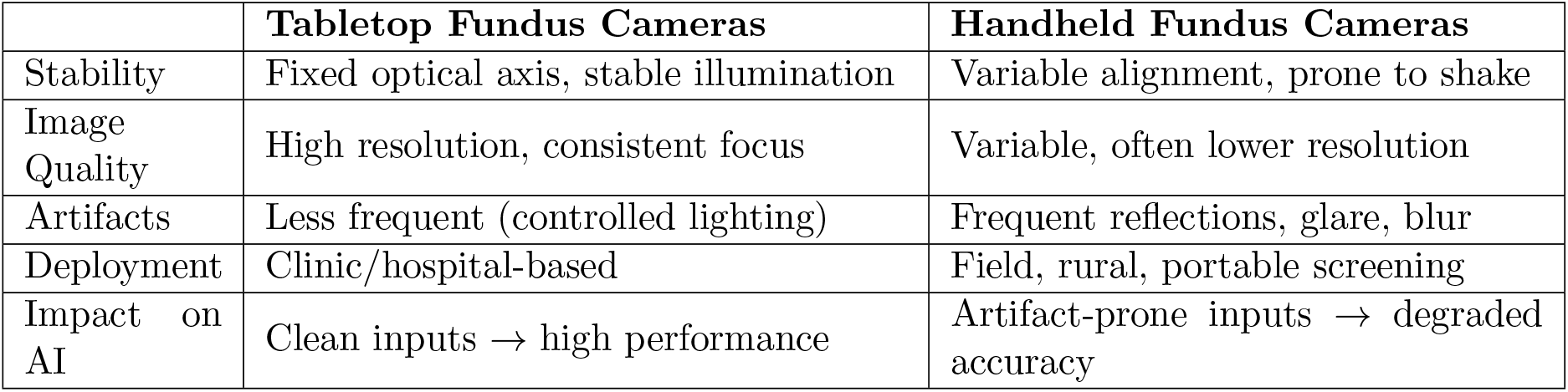
Comparison of imaging modalities and their artifact profiles.

|  | **Tabletop Fundus Cameras** | **Handheld Fundus Cameras** |
| --- | --- | --- |
| Stability | Fixed optical axis, stable illumination | Variable alignment, prone to shake |
| Image Quality | High resolution, consistent focus | Variable, often lower resolution |
| Artifacts | Less frequent (controlled lighting) | Frequent reflections, glare, blur |
| Deployment | Clinic/hospital-based | Field, rural, portable screening |
| Impact on AI | Clean inputs → high performance | Artifact-prone inputs → degraded accuracy |

## 4 Dataset and Augmentation Method

### 4.1 Original Dataset

The DDR dataset contains 13,673 high-resolution RGB retinal images labeled for DR severity and lesion segmentation. It is widely used for benchmarking.

### 4.2 Artifact Patch Extraction

We cropped artifact patches from anonymized images captured in handheld settings. Only optical artifact regions were retained, ensuring privacy.

### 4.3 Overlay Procedure

- Randomly select 1–32 patches per image.
- Resize each to 10–20% of retinal diameter.
- Apply circular feathered masks.
- Blend with OpenCV `seamlessClone` for natural integration.

## 5 Dataset Composition

The DDR-Augmented-Artifacts dataset includes:

- Augmented RGB images with reflection artifacts.
- Augmentation scripts for extensibility.
- Baseline ResU-Net model for artifact removal.

**Figure 2.**
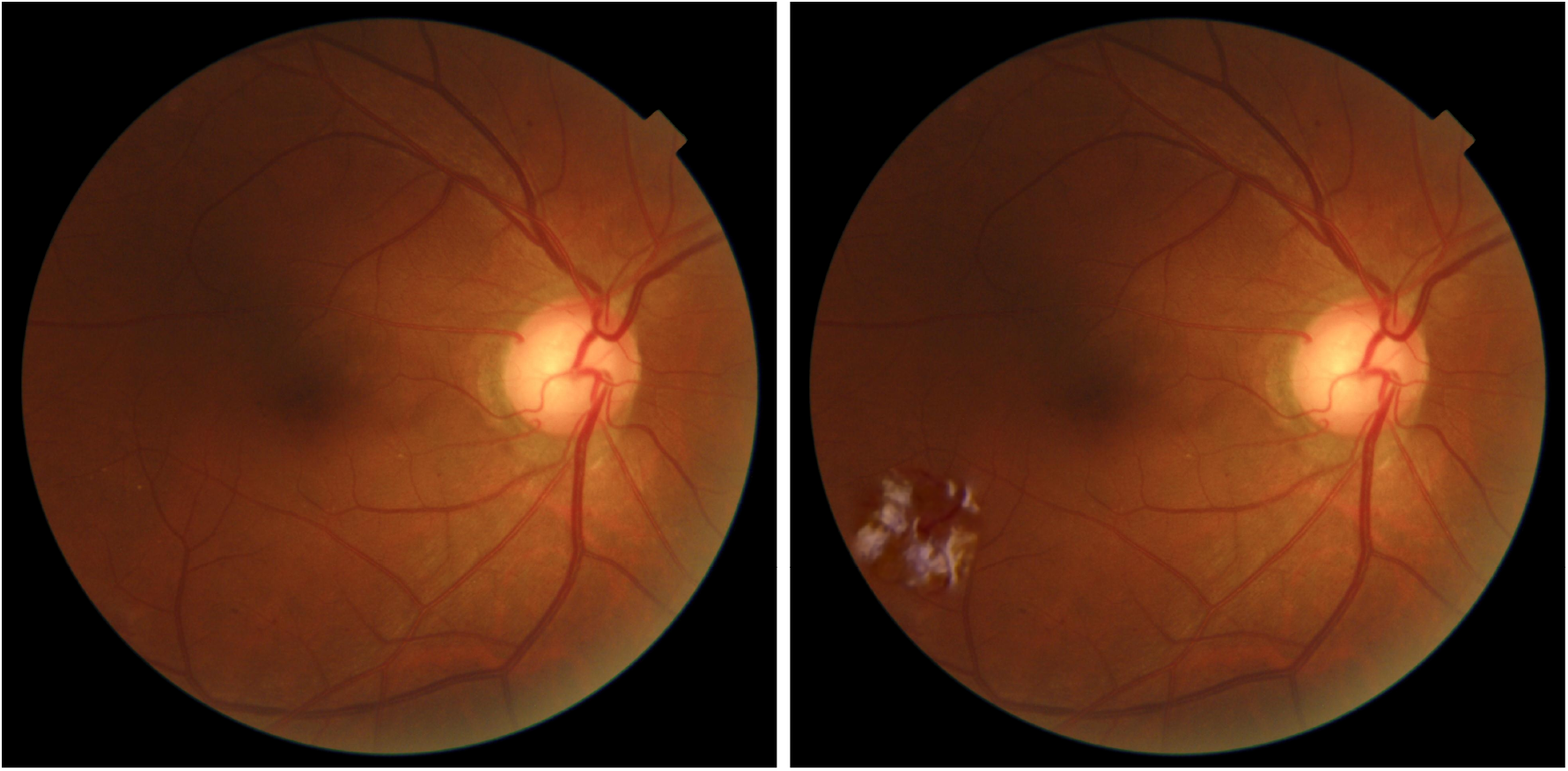
Example of artifact augmentation. Left: original DDR image. Right: augmented with synthetic reflection artifacts.

## 6 Demonstration: Artifact Removal Model

We trained a ResU-Net (residual U-Net) as a proof-of-concept. Training used:

- Loss: MSE + SSIM + perceptual loss.
- Input: artifact-augmented DDR.
- Output: cleaned image.

## 7 Why This Dataset Matters

Most studies either:

- Exclude noisy images (e.g., the NHS rejects ungradable inputs, which necessitates costly retakes).
- Use exclusively controlled tabletop data (e.g., IDx-DR, EyeArt). Our contribution:
- Provides controlled, reproducible artifact-rich data.
- Enables benchmarking robustness under realistic conditions.
- Encourages development of AI systems deployable in handheld-based screening.

**Figure 3.**
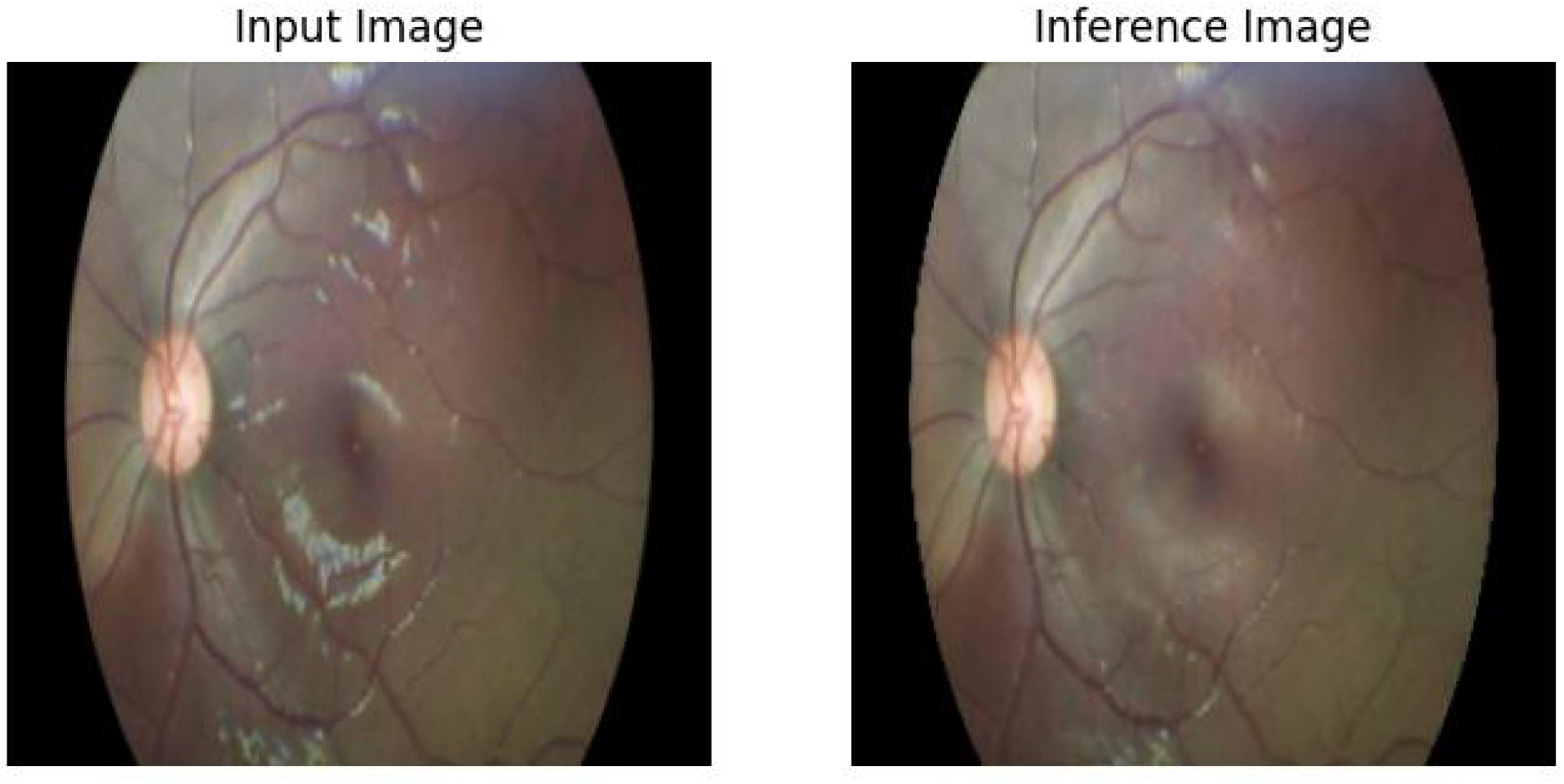
Comparison of input and output images. The left image shows the input with artifacts, and the right image shows the corresponding output produced by our ResU-net model.

## 8 Conclusion

We release DDR-Augmented-Artifacts, the first openly available dataset simulating reflection artifacts in DR fundus images. This bridges the gap between clean research datasets and noisy field conditions. Future work includes extending to blur and underexposure artifacts. By training and testing on such data, AI models will be more robust, clinically trustworthy, and deployable in real-world screening.

## Data Availability

The dataset,
augmentation scripts, and a sample demonstration model are publicly available at:
GitHub: https://github.com/Shubham2376G/DR_Artifacts
Hugging Face: https://huggingface.co/datasets/shubham212/DR_Artifacts
The original DDR dataset used in this study is publicly available at https://github.com/nkicsl/DDR-dataset

https://github.com/Shubham2376G/DR_Artifacts

https://huggingface.co/datasets/shubham212/DR_Artifacts

## Acknowledgements

We thank Tao Li et al. for the DDR dataset, and acknowledge prior contributions in DR screening datasets and algorithms including Gulshan et al., and Ting et al.

